# Diagnostic Utility of TRPS1 Immunohistochemistry in Primary and Metastatic Breast Carcinoma with Special Emphasis on Spindle Cell Neoplasms of the Breast

**DOI:** 10.1101/2025.11.12.25340091

**Authors:** Saroja Devi Geetha, Dongling Wu, Sudarshana Roychoudhury, Silvat Sheikh-Fayyaz

## Abstract

**Introduction:** Breast cancer, a major contributor to cancer-related deaths in females, presents significant challenges in classification and diagnosis, particularly triple negative breast cancers (TNBC). This study explores the utility of Trichorhinophalangeal syndrome type 1 (TRPS1) as a novel biomarker, comparing its efficacy with GATA3 in TNBC with a specific focus on spindle cell neoplasms.

**Methods:** Our pathology database was searched to identify breast carcinoma cases between January 2021 and July 2021. All cases of poorly differentiated breast carcinoma (primary and metastatic) and primary spindle cell neoplasms of breast confirmed on core biopsies, in women greater than 18 years were selected. Immunohistochemical staining for ER, PR, HER2, GATA3 and TRPS1 were interpreted by two pathologists. TRPS1 and GATA3 expression levels among different groups were analyzed.

**Results:** Our study cohort comprised of 60 cases (24 primary breast carcinomas, 22 metastatic breast carcinomas and 14 spindle cell lesions of the breast). Our study revealed TRPS1’s superior sensitivity in TNBC, with 84% positivity compared to 53% for GATA3. TRPS1 showed promise in diagnosing spindle cell neoplasms, identifying 6/14 (43%) of cases, while GATA3 yielded no positive results. The six spindle cell cases that were TRPS1 positive were one synovial sarcoma, two malignant phyllodes, one fibromatosis, one myofibroblastoma and one angiosarcoma.

**Conclusion:** TRPS1 holds substantial promise as a sensitive and novel diagnostic marker for TNBC in comparison to GATA3. The differential diagnostic utility of TRPS1 warrants careful consideration due to its positivity in other spindle cell neoplasms of the breast, which may present as differential diagnoses for metaplastic carcinoma.

**Highlights:**

- TRPS1 demonstrated superior sensitivity (84%) compared to GATA3 (53%) in identifying triple-negative breast carcinomas (TNBC), supporting its role as a promising diagnostic marker.
- TRPS1, unlike GATA3, showed positivity in 43% of spindle cell breast neoplasms, including malignant phyllodes, fibromatosis, myofibroblastoma, angiosarcoma, and synovial sarcoma, highlighting its broader expression spectrum.
- Pathologists should exercise caution in interpreting TRPS1 positivity within spindle cell lesions, as its expression may mimic metaplastic carcinoma, underscoring the need for careful correlation with morphology and other markers.

## Introduction

Breast cancer, a heterogeneous group of tumors, is the second most common leading cause of cancer mortality in the females.[1] Current classification methods rely on histology and hormone receptor expression, such as estrogen receptor (ER), progesterone receptor (PR), and ERBB2 receptor (HER2). Triple-negative breast cancer (TNBC), characterized by the absence of these receptors, comprises approximately 20% of cases and is known for its aggressive nature and high-grade histology.[2]

GATA binding protein 3 (GATA3) has been utilized alongside morphological analysis and other markers like gross cystic disease fluid protein 15 (GCDFP-15) and mammaglobin to confirm the breast origin. However, recent studies have revealed that GATA3’s sensitivity, especially in TNBC with high-grade histology, can drop as low as 40%.[3] Trichorhinophalangeal syndrome type 1 (TRPS1) has been shown to be a highly sensitive biomarker for breast cancer, including TNBC and metaplastic carcinoma, surpassing GATA3 in terms of sensitivity and specificity.[4]

Spindle cell carcinomas and fibromatosis-like metaplastic carcinomas are predominantly composed of spindle cells. The lack of an epithelial component poses challenges in differentiating it from other spindle lesions such as fibromatosis and malignant phyllodes. However, the role of TRPS1 as a biomarker for other spindle cell neoplasms of the breast, and its sensitivity and specificity in this context, remains poorly established. Thus, the objective of this study is to assess the utility of TRPS1, particularly its immunohistochemical expression, in TNBC and metastatic breast carcinomas, with a specific focus on spindle cell neoplasms of the breast.

## Methods

A retrospective search of our pathology database, Cerner Millenium was performed to identify breast carcinoma cases between January 2021 and July 2021. All cases of poorly differentiated breast carcinoma (primary and metastatic) and primary spindle cell neoplasms of breast confirmed on core biopsies, in women greater than 18 years were selected.

For each of these cases, one non-frozen, formalin-fixed, paraffin-embedded block with the largest portion of tumor was selected and tissue sections of 4um thickness were prepared for TRPS1 and GATA staining. Quality assurance measures were taken to examine these slides for folded, blurred, or obstructed morphologies. All cases had ER, PR and HER2 staining which was done at the time of initial diagnosis for patient care purposes.

Immunohistochemical staining for TRPS1 was performed with a rabbit polyclonal antibody (PA5-845874 from Invitrogen/ ThermoFisher, Waltham, MA) with a dilution of 1:500. Staining for GATA3 was done using a mouse monoclonal antibody (L50-823 from Cell Marque, Rocklin, CA), which is prediluted to 1.50 micrograms. The antibodies used during original diagnosis for hormonal immunoreactivity are ER antibody (Clone SP1 from Ventana), PR antibody (Clone 1E2), and HER-2 receptor antibody (Clone 4B5, Ventana).

IHC interpretation and scoring were done by two breast pathologists. ER/PR/HER2 scores were reported based on College of American Pathologists (CAP) and the American Society of Clinical Oncology (ASCO) guidelines. Accordingly, ER and PR were considered positive if nuclear staining was observed in more than 1% of tumor cells, while they were considered negative if nuclear staining was observed in less than 1% of tumor cells. HER-2/neu status was determined as negative if scored as 0 or 1+ (indicating incomplete, mild to moderate membrane staining in >10% of tumor cells), while it was considered positive if scored as 3+ (indicating complete, strong membrane staining in >10% of tumor cells). HER-2 /neu staining was reported equivocal if scored as 2+ and underwent chromogenic in situ hybridization (CISH) testing for further classification. Cases were considered as HER2amplified if copy number ≥ 6 per cell or HER2:CEP17 ratio ≥ 2.

For TRPS1 and GATA3, the percentage of nuclear positivity in tumor cells and the intensity of staining were assessed. Percentage of positive nuclear staining to total tumor cells were calculated and numerical values were assigned: 0:< 1%, 1: 1-50%, 2 > 50%. Numerical values were also assigned to the intensity of staining: 0 = negative; 1 =weak; 2 = moderate; 3 = strong. A composite score was calculated by multiplying these two numerical values. Based on the composite score, the overall staining pattern was divided into 3 groups: low:1; moderate:2-3; high:4-6. For the purpose of our study, we considered the cases demonstrating moderate and high intensity staining as positive. The TRPS1 and GATA3 expression levels (positive or negative) among different groups were compared. Fishers exact test was conducted among various subtypes of lesions to compare the IHC staining of TRPS1 and GATA3. A value of >0.05 was considered as statistically significant.

This study was approved by the Institutional Review Board (IRB) of the Human Research Protection Program licensing committee at Northwell Health. All guidelines and regulations were strictly followed in conducting the study.

## Results

Our study cohort comprised a total of 60 cases of which 24 were primary breast carcinomas, 22 metastatic breast carcinomas and 14 were spindle cell lesions of the breast. The neoplasms categorized under spindle cell morphology encompassed 5 myofibroblastomas, 3 fibromatosis, 2 angiosarcomas, 2 malignant phyllodes, 1 dermatofibrosarcoma protuberans (DFSP) and 1 synovial sarcoma.

Among the primary breast cancer cases, 4 were ER/PR+, 1 exhibited HER2+ and 19 were TNBC, categorized into 11 metaplastic and 8 non-metaplastic subtypes. Tumors within the metaplastic category consisted of cases with following differentiation: 4 spindle, 4 heterologous (3 chondroid and 1 osteoid) and 3 squamous. In the metastatic breast carcinoma group, 13 cases were ER/PR positive, 1 displayed HER2 positivity, and 8 were categorized as TNBC.

The expression of TRPS1 was observed in 3/4 (75%) of ER/PR+ primary breast cancer cases, while none of the HER2+ cases exhibited TRPS1 staining. In contrast, GATA3 demonstrated 100% positivity rates in both ER/PR+ and HER2+ cases. Within the TNBC cohort the TRPS1 positivity rates were higher than GATA3 positivity (83% vs 53%). [Table 1]

**Table 1.** TRPS1 and GATA3 immunohistochemical expression in primary and metastatic breast carcinoma and spindle cell lesions.

|  | Primary Breast Carcinoma |  |  |  |  |  |  |
| --- | --- | --- | --- | --- | --- | --- | --- |
| Marker | Subtype |  | Negative | Positive Low | Positive Intermediate | Positive High | Total |
| TRPS1 |  | ER/PR+ | - | - | 1 | 3 | 4 |
|  |  | HER2+ | - | - | 1 | - | 1 |
|  | TNBC | Metaplastic | 2 | - | 1 | 8 | 11 |
|  |  | Non-metaplastic | - | - | - | 8 | 8 |
|  |  | Other spindle cell neoplasms | 3 | 5 | 5 | 1 | 14 |
| GATA-3 |  | ER/PR+ | - | - | - | 4 | 4 |
|  |  | HER2+ | - | - | - | 1 | 1 |
|  | TNBC | Metaplastic | - | 5 | 4 | 2 | 11 |
|  |  | Non-metaplastic | 4 | - | - | 4 | 8 |
|  |  | Other spindle cell neoplasms | 11 | 3 | - | - | 14 |
|  | Metastatic Breast Carcinoma |  |  |  |  |  |  |
| TRPS1 | ER/PR+ |  | 1 | - | - | 12 | 13 |
|  | HER2+ |  | - | - | - | 1 | 1 |
|  | TNBC |  | 2 | - | 1 | 5 | 8 |
| GATA-3 | ER/PR+ |  | - | - | - | 13 | 13 |
|  | HER2+ |  | - | - | - | 1 | 1 |
|  | TNBC |  | 2 | - | - | 6 | 8 |

Among spindle cell neoplasms, 6/14 (43%) demonstrated TRPS1 positivity, with none of these cases showing GATA3 expression. The six TRPS1-positive cases included one synovial sarcoma, two malignant phyllodes tumors, one fibromatosis, one myofibroblastoma, and one angiosarcoma (Figure 1). Among these, only synovial sarcoma demonstrated a high staining score, while the other 5 cases demonstrated an intermediate score. Fishers exact test performed to compare the TRPS1 IHC with GATA3 IHC on spindle cell neoplasm was 0.007, which is statistically significant.

**Figure 1.**
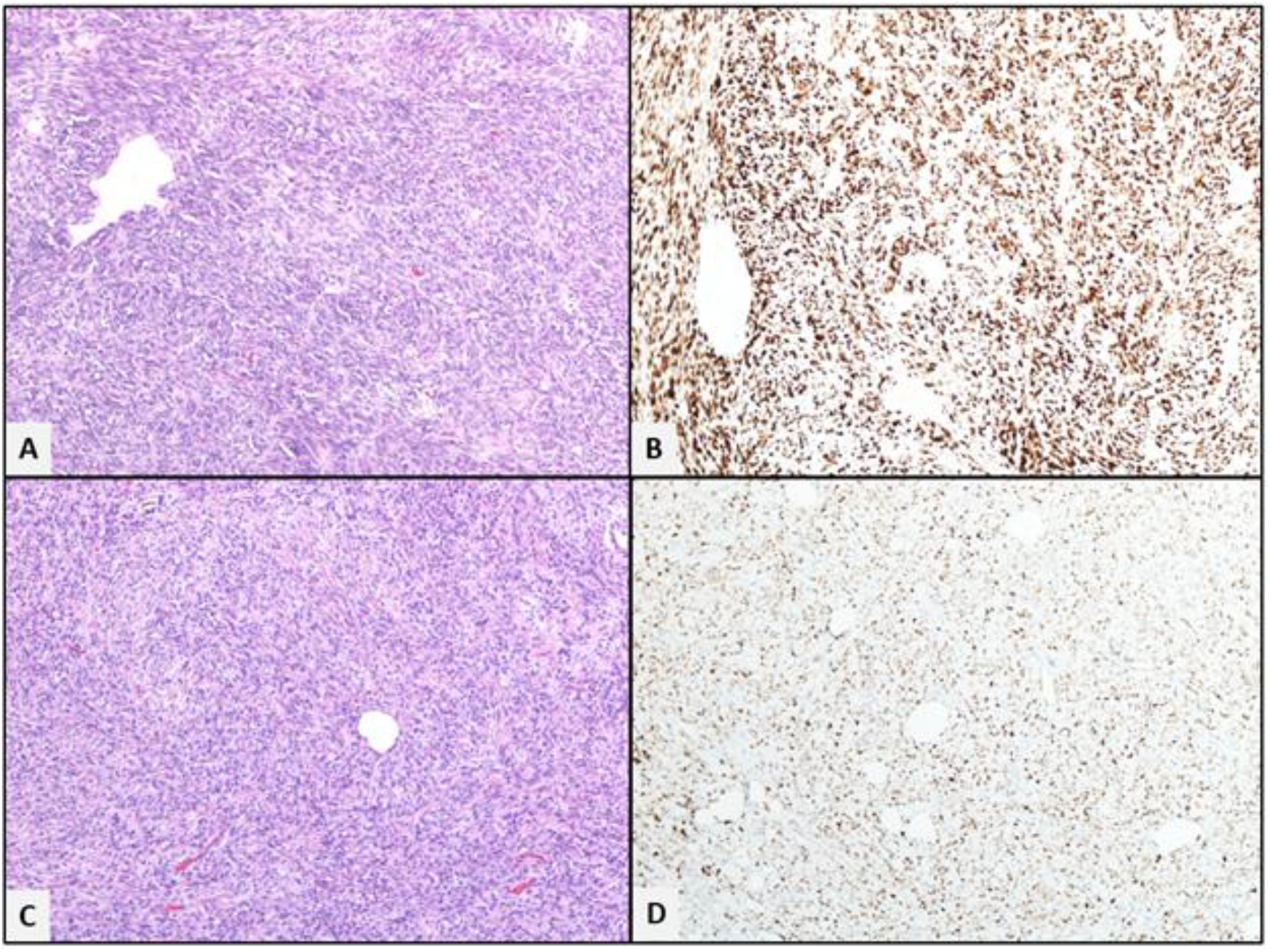
TRPS1 staining pattern in spindle cell neoplasm of breast. A: Synovial sarcoma (H& E, 10X). B: High positive TRPS1 staining pattern in synovial sarcoma (10 X) C: Myofibroblastoma (H& E, 10X) D: Intermediate positive TRPS1 staining in myofibroblastoma (10X)

Within the metastatic breast carcinomas, 12/13 (92%) ER/PR+ cases demonstrated TRPS1 staining, while all 13 cases demonstrated GATA3 positivity The sole HER2 positive case demonstrated both GATA3 and TRPS1 staining. TNBC demonstrated 6/8 (75%) positivity rates with both GATA3 and TRPS1.

## Discussion

Trichorhinophalangeal syndrome type 1 (TRPS-1), is a transcriptional regulator, belonging to the GATA family and is located on the long arm of chromosome 8q24. It has been known to be associated with an autosomal dominant malformation syndrome that results in craniofacial and skeletal abnormalities. [5] In 2005, TRPS1 was identified to be one of the most highly overexpressed genes in breast carcinomas.[6] Since then, TRPS-1 has been highly studied and has emerged to be a highly sensitive immunohistochemical marker in diagnosing breast cancers. It is found to have better rates at diagnosing triple negative breast cancers both in tissue sections and cytology when compared to already existing IHC markers such as GATA3 and SOX10.[7, 8]

Our study demonstrated that the TRPS1 staining in TNBCs had higher positivity rates than GATA3, thereby confirming existing studies showing that TRPS-1 is a more sensitive marker than GATA3 in breast cancers. The observation of lower specificity for TNBC has been consistent across multiple studies. Furthermore, we saw that a significant number of non-metaplastic spindle cell lesions that were all GATA3 negative, demonstrated TRPS1 positivity including synovial sarcoma, malignant phyllodes, fibromatosis, myofibroblastoma and angiosarcoma. These findings further support the existing literature. Study by Wang et al demonstrated a high proportion of malignant phyllodes tumor and sarcomas with osseous and chondroid differentiation in their cohort (95%) demonstrated strong TRPS-1staining as opposed to extramammary sarcomas.[9] Their study had 12 cases of post-radiation angiosarcoma none of which demonstrated TRPS-1 positivity. In our study cohort, one case of synovial sarcoma arising from the breast demonstrated TRPS1 staining. The expression of TRPS-1 in synovial sarcoma has been previously demonstrated by Cloutier et al.[10] Their study included primary, metastatic and recurrent tumors, with TRPS1 positivity in 86% of cases. However, they did not have a breast primary.

Therefore, when dealing with a breast core biopsy with a spindle cell morphology, including TRPS-1 in the IHC panel can help distinguish spindle cell lesions of the breast from the metaplastic lesions. Furthermore, in instances of angiosarcoma, TRPS-1 positivity supports a primary angiosarcoma rather than post-radiation angiosarcoma. When encountered with a metastatic spindle cell lesion, TRPS-1 staining in such instances could help point to a breast origin.

Despite initially being identified as a highly specific marker for breast tissue, subsequent investigations have demonstrated TRPS1 staining in tumors originating from Mullerian, lung, urothelial, salivary gland, gastrointestinal tract and bone and soft tissues.[7, 11-13]

Among tumors of Mullerian origin, it has been observed that serous neoplasms exhibit higher expression rates of TRPS1 compared to non-serous neoplasms.[7, 12] Notably, serous ovarian tumors are the most prevalent non-mammary metastases to the breast and axilla.[14] This adds an additional layer of complexity to the diagnostic applicability of TRPS1, as relying solely on TRPS1 expression may lead to misinterpretation of such cases as primary breast carcinomas.

TRPS1 expression has been observed in both primary and metastatic tumors of the salivary gland. [15, 16] Among the benign tumors, only pleomorphic adenoma exhibited TRPS1 expression. In contrast, TRPS1 expression in malignant tumors varied, with basal cell adenocarcinoma, polymorphous adenocarcinoma, secretory carcinomas, and adenoid cystic carcinomas demonstrating strong positivity. The similarities in architectural structure and cellular components between the breast and salivary gland make them prone to developing similar neoplasms. As a result, when confronted with a metastatic lesion involving either of these tumors, the use of TRPS1 as a diagnostic marker becomes highly challenging in discerning the primary origin.

One of the earlier studies demonstrated that osteosarcomas showed a TRPS1 positivity rate of 32% and that TRPS1 expression in osteosarcomas was associated with distant metastasis and reduced 3-year survival rates. [17] Similarly, synovial sarcomas have been found to demonstrate extremely high levels of TRPS1 expression (86%).[10]

A recent study demonstrated a notable prevalence (86%) of TRPS1 expression in sweat gland tumors (SGTs).[18] While it failed to discriminate between benign and malignant SGTs, it identified a positive TRPS1 staining pattern in tumors characterized by polygonal cells forming islands or nodules, such as hidradenomas, hidradenocarcinomas, poroma, porocarcinomas. Conversely, tumors with small ducts or strands of cells, such as microcystic adnexal carcinoma exhibited negative TRPS1 staining. The observed variation in staining patterns among different types of sweat gland tumors indicates the likelihood of different cellular origins or divergent pathways of cellular differentiation. These findings present an opportunity for future research and the potential utilization of TRPS1 as a diagnostic tool in sweat gland tumors.

This study has several limitations. The relatively small sample size, particularly of spindle cell lesions, may restrict the generalizability of the findings. Being a retrospective study conducted at a single institution, it is also subject to selection bias and variations in tissue fixation, processing, and staining, which could influence immunohistochemical results. Only one antibody clone for TRPS1 was utilized, and potential differences in clone performance or staining platforms were not explored, which may affect reproducibility in other settings.

## Conclusion

In this study, we have discovered that TRPS1 holds substantial promise as a sensitive and novel diagnostic marker for TNBC in comparison to GATA3. The differential diagnostic utility of TRPS1, however, warrants careful consideration due to its potential positivity in other spindle cell neoplasms of the breast, which may present as differential diagnoses for metaplastic carcinoma. Consequently, it is imperative for pathologists to maintain a high level of awareness regarding this characteristic while engaging in the interpretation of spindle cell lesions observed in breast tissue samples. Such awareness is crucial in ensuring accurate and precise diagnoses, thereby facilitating appropriate treatment strategies and patient management. Further investigation and validation studies are warranted to fully elucidate the diagnostic potential and clinical implications of TRPS1 in the context of breast spindle cell lesions.

## Data Availability

All data produced in the present study are available upon reasonable request to the authors

## Notes

### Competing Interest Statement

The authors have declared no competing interest.

### Funding Statement

This study did not receive any funding

### Author Declarations

Ethics committee/IRB of Northwell Health gave ethical approval for this work

## References

[1] C. f. D. C. a. Prevention, “An Update on Cancer Deaths in the United States. Atlanta, GA: US Department of Health and Human Services, Centers for Disease Control and Prevention, Division of Cancer Prevention and Control,” ed, 2022.

[2] H. Yao et al., “Triple-negative breast cancer: is there a treatment on the horizon?,” (in eng), Oncotarget, vol. 8, no. 1, pp. 1913–1924, Jan 03 2017, doi: 10.18632/oncotarget.12284.

[3] L. Huo et al., “GATA-binding protein 3 enhances the utility of gross cystic disease fluid protein-15 and mammaglobin A in triple-negative breast cancer by immunohistochemistry,” (in eng), Histopathology, vol. 67, no. 2, pp. 245–54, Aug 2015, doi: 10.1111/his.12645.

[4] B. Parkinson, W. Chen, T. Shen, A. V. Parwani, and Z. Li, “TRPS1 Expression in Breast Carcinomas: Focusing on Metaplastic Breast Carcinomas,” (in eng), Am J Surg Pathol, vol. 46, no. 3, pp. 415–423, Mar 01 2022, doi: 10.1097/PAS.0000000000001824.

[5] P. Momeni et al., “Mutations in a new gene, encoding a zinc-finger protein, cause tricho-rhino-phalangeal syndrome type I,” (in eng), Nat Genet, vol. 24, no. 1, pp. 71–4, Jan 2000, doi: 10.1038/71717.

[6] L. Radvanyi et al., “The gene associated with trichorhinophalangeal syndrome in humans is overexpressed in breast cancer,” (in eng), Proc Natl Acad Sci U S A, vol. 102, no. 31, pp. 11005–10, Aug 02 2005, doi: 10.1073/pnas.0500904102.

[7] D. Ai et al., “TRPS1: a highly sensitive and specific marker for breast carcinoma, especially for triple-negative breast cancer,” (in eng), Mod Pathol, vol. 34, no. 4, pp. 710–719, Apr 2021, doi: 10.1038/s41379-020-00692-8.

[8] M. Abdelwahed et al., “Utility of TRPS-1 immunohistochemistry in diagnosis of metastatic breast carcinoma in cytology specimens,” (in eng), J Am Soc Cytopathol, vol. 11, no. 6, pp. 345–351, 2022, doi: 10.1016/j.jasc.2022.06.007.

[9] J. Wang et al., “Expression of TRPS1 in phyllodes tumor and sarcoma of the breast,” (in eng), Hum Pathol, vol. 121, pp. 73–80, Mar 2022, doi: 10.1016/j.humpath.2022.01.002.

[10] J. M. Cloutier, D. R. Ingram, K. Wani, A. J. Lazar, and W. L. Wang, “Frequent TRPS1 expression in synovial sarcoma is associated with SS18-SSX fusion oncoprotein activity,” (in eng), Hum Pathol, vol. 130, pp. 88–94, Dec 2022, doi: 10.1016/j.humpath.2022.09.006.

[11] M. Wang, K. Stendahl, G. Cai, A. Adeniran, M. Harigopal, and S. M. Gilani, “Evaluation of TRPS1 Expression in Pleural Effusion Cytology Specimens With Metastatic Breast Carcinoma,” (in eng), Am J Clin Pathol, vol. 158, no. 3, pp. 416–425, Sep 02 2022, doi: 10.1093/ajcp/aqac066.

[12] S. D. Geetha et al., “TRPS1 Function Beyond Breast: A Retrospective Immunohistochemical Study on Non-breast Cytology Specimens,” Journal of the American Society of Cytopathology, vol. 12, no. 5, pp. S7–S8, 2023/09/01/ 2023, doi: 10.1016/j.jasc.2023.07.014.

[13] A. Ali et al., “TRPS1 function beyond breast: A retrospective immunohistochemical study on non-breast cytology specimens,” (in eng), Diagn Cytopathol, vol. 52, no. 9, pp. 499–504, Sep 2024, doi: 10.1002/dc.25359.

[14] D. F. DeLair, A. D. Corben, J. P. Catalano, C. E. Vallejo, E. Brogi, and L. K. Tan, “Non-mammary metastases to the breast and axilla: a study of 85 cases,” (in eng), Mod Pathol, vol. 26, no. 3, pp. 343–9, Mar 2013, doi: 10.1038/modpathol.2012.191.

[15] C. G.-F. Youley Tjendra, Oleksandr Kryvenko, Jaylou Velez Torres, “TRPS1 Immunohistochemical Expression in Salivary Gland Tumors: A Pilot Study,” presented at the USCAP, New Orleans, 2023.

[16] S. D. Geetha et al., “TRPS1 Immunohistochemistry in Salivary Gland Neoplasms: Analysis on Cytology Cell Blocks and Surgical Follow-Up Correlation,” (in eng), Diagn Cytopathol, vol. 53, no. 3, pp. 121–126, Mar 2025, doi: 10.1002/dc.25428.

[17] Z. Li, M. Jia, X. Wu, J. Cui, A. Pan, and L. Li, “Overexpression of Trps1 contributes to tumor angiogenesis and poor prognosis of human osteosarcoma,” (in eng), Diagn Pathol, vol. 10, p. 167, Sep 17 2015, doi: 10.1186/s13000-015-0401-2.

[18] H. B. Zengin, C. M. Bui, K. Rybski, T. Pukhalskaya, B. Yildiz, and B. R. Smoller, “TRPS1 Is Differentially Expressed in a Variety of Malignant and Benign Cutaneous Sweat Gland Neoplasms,” (in eng), Dermatopathology (Basel), vol. 10, no. 1, pp. 75–85, Feb 02 2023, doi: 10.3390/dermatopathology10010011.

